# 13 cis retinoic acid improved the outcomes of COVID-19 patients. A randomized clinical trial

**DOI:** 10.1101/2022.03.05.22271959

**Authors:** Mahmoud Elkazzaz, Yousry Esam-Eldin Abo-Amer, Amr Ahmed, Tamer Haydara

## Abstract

The COVID-19 pandemic caused by severe acute respiratory syndrome coronavirus 2 (SARS-CoV-2) has infected over 100 million people causing over 2.4 million deaths over the world, and it is still expanding. Given the urgency of the COVID-19 pandemic, the clinical investigation of approved drugs is a promising alternative to find a timely effective treatment. In this randomized trial, we investigated the activity of both oral and aerosolized 13 cis retinoic acid in the treatment of SARS-COV-2 added to standard of care treatment in patients with COVID-19 versus standard of care treatment alone. This was a randomized controlled trial conducted at Kafrelsheikh University’s Quarantine Hospitals, Egypt. After obtaining informed consent, forty patients with a confirmed diagnosis of COVID-19 were enrolled in the study. They were randomly assigned to one of two groups: Group I; 20 patients received aerosolized and oral 13 cis retinoic acid plus standard of care treatment (13 cis RA group) and Group II; 20 patients received only standard care treatment as a control group. The two groups were age and gender matched. There was no statistically significant difference between them in any of the baseline characteristics or laboratory parameters. The results showed that there was a high significant difference between the two groups regarding intensive care unit (ICU) admission, mortality and improvement (P<0.05). Only 10.52 % of patients in the 13 cis retinoic acid group needed ICU admission compared to 28.57 % in the control arm. There was no mortality in the 13 cis retinoic acid group, whereas about 14.35% were died in the group II. All patients who received 13 cis retinoic acid noticed a high improvement (P<0.001), and the mean value for clinical improvement was 16.3±4.5 days. There was no significant difference regarding the laboratory parameters before and after 14 days of treatment in the group of patients received the standard of care treatment (P=0.66). Univariate logistic regression analysis showed overall mortality was significantly related to the patient’s age, serum ferritin, C-reactive protein, oxygen saturation, the presence of diabetes mellitus, obesity, and abdominal pain. We conclude that 13 cis retinoic acid is a promising drug in the treatment of patients with COVID-19 infection, when added to the standard of care treatment.

## Introduction

An outbreak of novel coronavirus pneumonia is ongoing, called COVID-19, was first identified in Wuhan, Hubei province, China at the end of 2019 (1, 2). The Chinese Center for Disease Control and Prevention (CDC) initially reported that the cause of this disease is severe acute respiratory syndrome coronavirus 2 (SARS-CoV-2), then it is founded to be a novel coronavirus (3). Now, this disease is designated by the World Health Organization (WHO) as COVID-19, which rapidly spreads to other cities of China, and has become a public health emergency of international concern (PHEIC). The clinical manifestations of COVID-19 are cough, fever, muscle pain, diarrhea, and pneumonia, and can cause death in severe cases (2). China has reported 72,435 cases of confirmed COVID-19 and 1,867 fatalities through February 18, 2020 (4). The entry of COVID-19 into respiratory epithelial cells depends on its spike protein and its attachment to the common receptors, human angiotensin-converting enzyme 2 (ACE2) receptors. The spike (S) protein of COVID-19 mainly, responsible for viral invasion and entry into lung cells. Viral entry depends on binding of the surface unit, S1, of the S protein to a cellular receptor, which facilitates viral binding to the surface of lung cells. SARS-S engages ACE2 receptors as the entry receptors and uses the cellular transmembrane protease, serine 2 (TMPRSS2) for S protein priming (4). The SARS-S/ACE2 interface has been elucidated at the atomic level, and the efficiency of ACE2 usage was found to be a key determinant of COVID-19 transmissibility (5). As the effective immune response in viral infections depends on the activation of cytotoxic T cells that can clear infection by killing virus-invaded cells, boosting the numbers and function of T cells in COVID-19 patients are needed for successful recovery. CD8 T cells produce very important and effective factors, such as Interferon gamma (IFN-g) and granzyme, to clear MERS-CoV (6). Also, low circulating lymphocyte counts in 82.2% of COVID-19 cases were showed by a recent study (7). A COVID-19 infects macrophages, and after that macrophages present COVID-antigens to T helper (Th0) cells. This process leads to T cell stimulation and activation, including the production of cytokines associated with the different T cell subsets (i.e., Th1, Th2, and Th17), followed by a huge release of cytokines for immune response amplification. The continued secretion of these immune factors due to viral persistence has a negative and unwanted effect on Th0 and cluster of differentiation 8 (CD8) T cell activation by inhibiting IL12 and IFN-g production (8). However, CD8 T cells produce very effective mediators, such as IFN-g and granzyme, to clear COVID-19. It is still unclear whether long-term or short-term protective antibodies are produced during neutralizing antibody production against COVID-19 (2). Attachment of COVID-19 to ACE2 on the host cell through S protein leads to the appearance of viral genomic ribonucleic acid (RNA) in the cytoplasm. An immune response to double-strand RNA (ds RNA) can be partially generated during COVID-19 replication. Toll-like receptor 3 (TLR-3) sensitized by dsRNA and cascades of signaling pathways (IRFs and NF-kB activation) are activated to produce type I IFNs and proinflammatory cytokines. Type I Interferon (IFNs) is important to support the release of antiviral proteins for the protection of uninfected cells. Accessory proteins of COVID-19 can interfere with TLR-3 signaling and bind the dsRNA of COVID-19 during replication to inhibit TLR-3 induction and activation and evade the immune response. Cell-virus interactions lead to high production of immune mediators. The secretion of large quantities of chemokines and cytokines is promoted in infected cells in response to MERS-CoV infection. These chemokines and cytokines in turn recruit lymphocytes and leukocytes to the site of infection (8). In a recent study of 522 COVID patients and 40 healthy controls from two hospitals in Wuhan, China showed a negative correlation between T cells numbers and the serum IL6, IL-10, and TNF-a concentration, on the other hand, patients in decline period showing reduced IL-6, IL-10, and TNF-a concentrations and restored T cell counts (9). Significantly higher levels of the exhausted marker programmed cell death protein (PD-1) in comparison to health controls, were expressed by T cells from COVID-19 patients. Moreover, as the patients progressed from prodromal to overtly symptomatic stages, it was reported that increasing PD-1 and T-cell immunoglobulin and mucin-domain containing-3 (Tim-3) expression on T cells, which further indicates T cell exhaustion. T cell exhaustion is a progressive loss of effector function due to prolonged antigenic stimulation, which is a characteristic of chronic infections (9). COVID-19 is World’s biggest challenge since World War II, says United Nations (UN) Secretary General. The emergence of coronavirus disease-2019 (COVID-19) pandemic and its catastrophic consequences resulted in extreme demand for treatment. This article investigates the potential effect of 13 cis retinoic acid, a potential metabolite of Vitamin A in the treatment of SARS-CoV-2 owing to its ability to downregulate angiotensin Converting Enzyme (ACE2) receptor and angiotensin II expression and its impact on immunity, androgen receptors, inflammatory IL-6, thrombin and respiratory diseases. Moreover, our recent molecular docking based analysis showed that spike protein Receptor Binding Domain (RDB) of COVID-19 strongly and efficiently binds to STRA6 receptor, definitely to the RDB vital residues of RBP-binding motif located in STRA6 receptor(10). STRA6 receptor expressed in many organs and immune cells, upregulated by retinoic acid 6 (STRA6), which was the first protein to be identified in a novel category of proteins, cytokine signaling transporters, due to its ability to function as both a cell surface receptor and a membrane protein that binds to retinol binding protein facilitating cellular uptake of retinol (10). The primary ligand of STRA6 (vitamin A/retinol) was shown to be drastically reduced during COVID-19 infection, which support the role of retinoic acid in COVID-19 treatment (10).

## Methods

### Study Design and Patients

We conducted a randomized interventional comparative trial a 1:1 ratio randomized (Two groups) using a computerized random number generator using simple randomization with an equal allocation ratio, comparing and investigating the efficacy of aerosolized and oral 13 cis retinoic acid plus Standard Therapy vs Standard Therapy including for treating COVID-19 among hospitalized adults with respiratory illness admitted to Kafrelsheikh University’s Quarantine Hospitals with suspected or confirmed COVID-19 in the period between June and August 2020. The trial was limited to include only laboratory-confirmed cases (Adult patients with COVID-2019 infection confirmed by PCR; Absolute value of lymphocytes < 1.0x 109/L). According to WHO interim guidelines released on March 13, 2020. Adults (aged ≥18 years) who had been hospitalized for less than 48 hours, patients were classified in to; **i) Mild cases**: represented patients with uncomplicated upper respiratory tract viral infection, **ii) moderate cases:** represented patients with pneumonia but without need for supplemental oxygen; **iii) Severe respiratory failure:** within 48 hours and requires admission to ICU and severe respiratory failure :was defined as PaO2/FiO2 < 200 mmHg and was supported by positive pressure mechanical ventilation (including non-invasive and invasive mechanical ventilation, PEEP>5cmH2O). This clinical trial was approved by the Research Ethics Committee of the Faculty of Medicine, Kafrelsheikh University. Informed and signed consent was obtained from patients or legally authorized representatives. The study was registered on clinicaltrials.gov with Clinical Trials.gov Identifier NCT04353180, It worth mentioning that this clinical trial of aerosolized 13 cis retinoic acid therapy was initially submitted on April 10, 2020, met QC Criteria on April 17, 2020, and was first published on April 20, 2020 (https://clinicaltrials.gov/ct2/keydates/NCT04353180).

In this study, a total of forty patients were included, and divided randomly in two groups as following: **Group I;**The 13 cis retinoic acid : This group included 20 patients who received aerosolized 13 cis retinoic acid for 14 days divided doses increased gradually in concentration and oral 13 cis retinoic acid with constant concentration (one dose increases froms 0.2 mg/kg/day to 4 mg/kg/day as inhaled 13 cis retinoic acid therapy) plus oral 13 cis retinoic acid 20 mg/day added to the standard therapy adopted according to treatment protocol by the national MOH for 14 days. **Group II;** This group included 20 patients who received only the standard therapy adopted according to treatment protocol by the national MOH for 14 days.

The Egyptian Ministry of Health (MOH) adopted a standard of care treatment protocol for COVID-19 patients. It included (Paracetamol 500 mg /6h, Hydroxychloroquine 500 mg/ 12h, Oseltamivir 150 mg /12 h for 5 days, Azithromycin 1 gm first day then 500 mg/day for 1st line or Clarithromycin 500 mg/12 h for 7-14 days, Ascorbic acid 500 mg/12 h and Cyanocobalamin IV once daily plus Lopinavir 400mg/Ritonavir 100 mg caps 2 capsules twice daily and invasive mechanical ventilation with hydrocortisone for severe cases if PaO2 < 60 mmHg, O2 saturation < 90% despite oxygen or noninvasive ventilation, progressive hypercapnia, respiratory acidosis (pH < 7.3), and progressive or refractory septic shock.

The history of all patients and also the appropriate clinical examination were done prior to the treatments. All the participants were subjected to thorough history taking and full clinical examination including age, gender, weight and height measurements, and calculation of body mass index (BMI); medication history; and investigations in the form of complete blood picture and computed tomography of the chest (CT chest).

#### Statistical analysis

Data were analyzed statistically using R programming, and were expressed the mean ± standard deviation (SD) or percentage (%). For normality investigation, the Shapiro–Wilks test was used for numerical data, and the Chi-square test (χ^2^) was used for categorical data. The means of samples with normal distribution and of sufficient size were compared by Student’s t test and one-way ANOVA. Differences between abnormal scatterings and the medians were compared by Mann-Whitney U and Kruskal-Wallis test. A p value < 0.05 was considered significant.

## Results

The clinical examinations of all patients in two groups were done, and the results showed that, Persistent fever was found in 26.3 %, intermittent fever was present in 37%, cough was present in 68.5 %, anosmia was found in 32 %, Abdominal pain and diarrhea was present in 22% of the included patients. With regards to oxygen saturation, Oxygen saturation between 95 and 90 was present in 52.63%, 90–85 in 15.7%, and less than 85 in 31.57% of all the participants. The computed tomography chest scans were normal in 31.57%, ground-glass opacities in 21.05%, confluent opacities in 15.5%, consolidation in 15.78%, and extensive consolidation in 15.7%. **(Table1)** showed that, there were no significant difference between both groups regarding all laboratory parameters Before treatment. The results revealed that in the first group of patients who received 13 cis retinoic acid, there was a highly significance difference before and after 14 days regarding laboratory parameters (**Table 2)**. On the other hand, in the patients group who received standard of care treatment, there was no significance difference after 14 days of treatment compared to at the start of treatment (**Table 3)**. The results of laboratory examination after 14 days of treatments revealed a high significance difference between the two groups **(Table 4)**.

**Table(1):** Laboratory parameters of both groups Before treatment.

| Character | Group 1(mean ±SD)<br>(Before14 day) | Group2 (mean ±SD)<br>(Before 14 day) | p-value |
| --- | --- | --- | --- |
| Lymphocytic count cells | 0.77 ± 0.10 | 0.78 ± 0.11 | 0.9 |
| crp | 101.74± 24.83 | 101.95± 27.94 | 0.8 |
| Ddimer.mg.L. | 0.73± 0.16 | 0.70 ± 0.14 | 0.63 |
| thrombin..IU.ml | 82.95± 5.95 | 82.05 ± 8.27 | 0.69 |
| ferritin | 741.16± 115.80 | 749.14 ±101.75 | 0.81 |
| thrombin.time | 9.57 ± 3.69 | 11.14 ± 4.68 | 0.24 |
| cholesterol..mmol.l. | 3.55± 0.19 | 3.56 ± 0.18 | 0.93 |
| testosterone | 8.00± 0.50 | 8.00 ± 0.39 | 0.97 |
| DH.testosterone | 1.74± 0.20 | 1.83 ± 0.30 | 0.246 |
| IgA.mg.dL | 111.32± 14.72 | 106.14 ± 17.90 | 0.34 |
| IL6.pg..l | 22.42± 11.32 | 20.46± 7.09 | 0.52 |
| IFN.Alpha.pg.ml | 17.95± 5.82 | 20. 05 ± 5.21 | 0.238 |
| Soluble.ACE2.U.L | 91.74± 26.27 | 94.67 ± 25.97 | 0.725 |
| Ag.il..pg.mL | 56.84± 12.69 | 59.86 ± 10.73 | 0.425 |

**Table (2):** Laboratory parameters of 13 cis retinoic acid group Before and after 14 days of treatment.

| Character | Before 14 day<br>(mean ±SD) | After 14 day<br>(mean ±SD) | p-value |
| --- | --- | --- | --- |
| Lymphocytic count cells | 0.77 ± 0.10 | 1.32± 0.21 | <0.001 |
| CRP | 101.74± 24.83 | 8.47 ± 16.53 | <0.001 |
| D-dimer (mg/L) | 0.73± 0.16 | 0.52 ± 0.16 | <0.001 |
| Thrombin (IU/ml) | 82.95± 5.95 | 54.00 ± 13.99 | <0.001 |
| Ferritin | 741.16± 115.80 | 339.32± 62.13 | <0.001 |
| Cholesterol (mmol/l) | 3.55± 0.19 | 5.08 ± 0.44 | <0.001 |
| DH.testosterone | 1.74± 0.20 | 1.54 ±0.20 | 0.004 |
| IgA (mg/dL) | 111.32± 14.72 | 119.58± 17.97 | 0.04 |
| IL6 (pg/l) | 22.42± 11.32 | 9.11 ± 4.40 | <0.001 |
| IFN-Alpha (pg/ml) | 17.95± 5.82 | 22.37± 6.52 | 0.03 |
| Soluble ACE2 (U/L) | 91.74± 26.27 | 58.42 ± 22.12 | <0.001 |
| Angiotensin II (pg/mL) | 56.84± 12.69 | 39.26 ± 9.39 | <0.001 |

**Table (3):** Laboratory parameters of the standard of care treatment group Before and after 14 days of treatment.

| Character | Before 14 day<br>(mean ±SD) | After 14 day)<br>(mean ±SD) | p-value |
| --- | --- | --- | --- |
| Lymphocytic count cells | 0.78 ± 0.11 | 1.01± 0.24 | 0.3 |
| CRP | 101.95± 27.94 | 99.90 ± 22.23 | 0.6 |
| D-dimer (mg/L) | 0.70 ± 0.14 | 0.68 ± 0.21 | 0.66 |
| Thrombin (IU/ml) | 82.05 ± 8.27 | 83.95 ± 5.74 | 0.76 |
| Ferritin | 749.14 ±101.75 | 632.62 ± 250.00 | 0.07 |
| Cholesterol (mmol/l) | 3.56 ± 0.18 | 3.7 ± 0.17 | 0.6 |
| DH.testosterone | 1.83 ± 0.30 | 5.35 ± 16.65 | 0.35 |
| IgA (mg/dL) | 106.14 ± 17.90 | 119.05± 29.12 | 0.09 |
| IL6 (pg/l) | 20.46± 7.09 | 16.57 ± 8.63 | 0.12 |
| IFN-Alpha (pg/ml) | 20.05 ± 5.21 | 17.43 ± 5.68 | 1 |
| Soluble ACE2 (U/L) | 94.67 ± 25.97 | 107.48 ± 76.94 | 0.6 |
| Angiotensin II (pg/mL) | 59.86 ± 10.73 | 53.29 ± 9.76 | 1 |

**Table (4):** Laboratory parameters between the two groups after 14 days of treatment.

| Character | Group 1(mean ±SD)<br>(After 14 day) | Group2 (mean ±SD)<br>(After 14 day) | p-value |
| --- | --- | --- | --- |
| Lymphocytic count cells | 1.32± 0.21 | 0.99 ± 0.16 | <0.001 |
| CRP | 8.47 ± 16.53 | 34.1 ±52.11 | 0.04 |
| D-dimer (mg.L) | 0.52 ± 0.16 | 0.63 ± 0.11 | 0.01 |
| Thrombin (IU/ml) | 54.00 ± 13.99 | 89.86 ± 6.92 | <0.001 |
| Ferritin | 339.32± 62.13 | 530.24±149.67 | <0.001 |
| Cholesterol (mmol/l) | 5.08 ± 0.44 | 3.91 ± 0.17 | <0.001 |
| DH.testosterone | 1.54 ±0.20 | 5.35 ± 16.65 | 0.3 |
| IgA (mg/dL) | 219.58± 53.97 | 126.14± 29.70 | <0.001 |
| IL6(pg/l) | 9.11 ± 4.40 | 13.76 ± 5.65 | <0.01 |
| IFN-Alpha (pg/ml) | 22.37± 6.52 | 13.43 ± 4.18 | 0.02 |
| Soluble ACE2 (U/L) | 58.42 ± 22.12 | 106.81±77.34 | 0.01 |
| Angiotensin II (pg/mL) | 39.26 ± 9.39 | 52.71±9.51 | <0.001 |

With regards to the clinical outcomes, in the 13 cis retinoic acid group, only 10.52 % of patients needed intensive care unit (ICU) admission compared to 28.57 % in the other group. In the 13 cis retinoic acid group, duration to achieve negative PCR was 13.36± 1.49 compared to 23.85±4 in the other group.

.The 13 cis retinoic acid group had a mean of 16.3±4.5 days to show clinical improvement and 21.57±4.97 days to hospital discharge, whereas the control group had a mean of 25.23±4.72 to clinical improvement and 33.04±2.72 to hospital discharge (P < 0.01). After 28 days, there was significant difference between the two groups regarding the clinical outcome (P < 0.05). Complete recovery was achieved in all cases (100%) of the 13 cis retinoic acid group, and there were no deaths in this group, While 52.38 % of patients who treated with standard of care treatment were recovered, 14.28 % mild, 4.76 % moderate 14.35% were died **(Table 5)**. By logistic regression, the overall mortality was not significantly associated with 13 cis retinoic acid therapy. On the other hand, it was significantly related to the C-reactive protein (P<0.001), D-dimer (P<0.001), ferritin (P<0.001), Oxygen saturation (P<0.05), obesity (P<0.001), the presence of diabetes mellitus (P<0.001), and abdominal pain and diarrhea (p<01) **(Table 6)**.

**Table (5):**
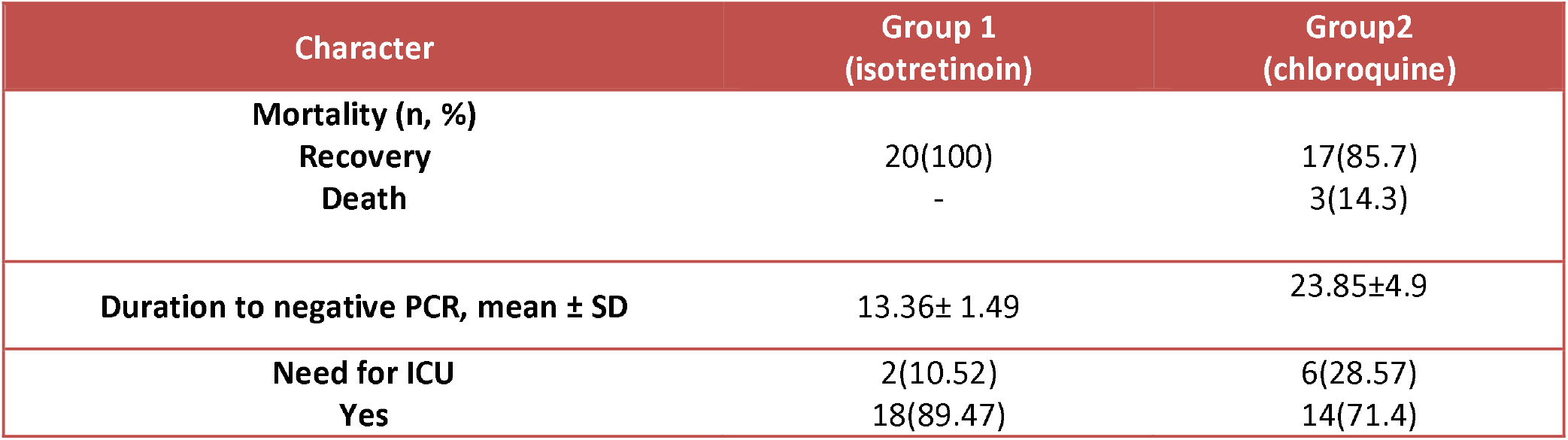
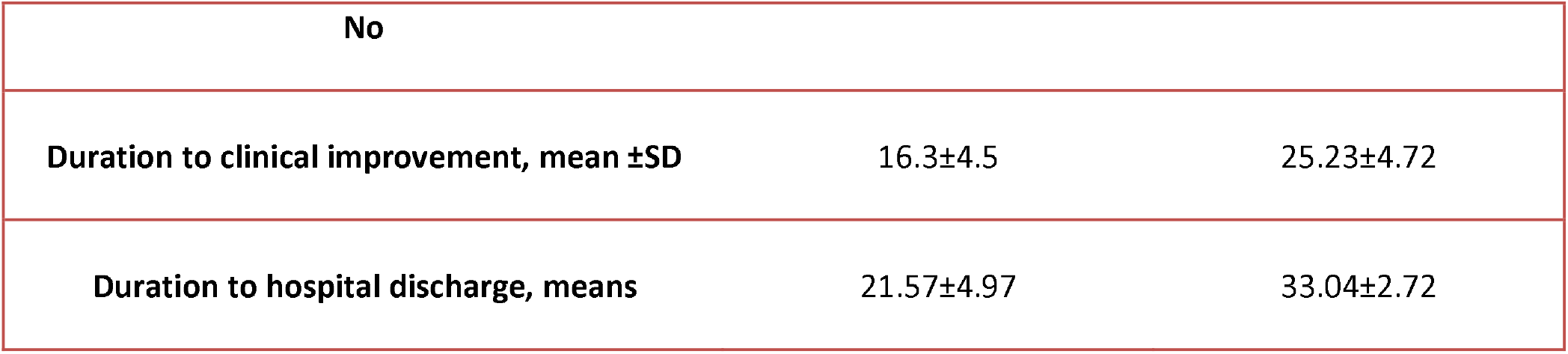
Clinical out course in both groups.

**Table (6):** Univariate regression of the potential risk factors of mortality.

| Variable | P value | OR | 95%CI |  |
| --- | --- | --- | --- | --- |
| Age | <0.001 | 1.53 | 1.33 | 1.50 |
| Gender | 0.3 | 1.2 | 0.82 | 1.77 |
| CRP | <0.001 | 1.0 | 1.0 | 1.0 |
| Ferritin | <0.001 | 1.0 | 1.0 | 1.0 |
| Diabetic | <0.001 | 0.54 | 0.45 | 0.65 |
| Obese | <0.001 | 1.53 | 1.22 | 1.92 |
| Cough | 0.2 | 1.13 | 0.87 | 1.48 |
| O2 saturation | <0.05 |  |  |  |
| 90-95 | 0.05 | 0.86 | 0.96 | 1.1 |
| 85-90 | <0.01 | 0.97 | 1.0 | 1.12 |
| >85 | 0.001 | 1.42 | 1.04 | 1.95 |
| Anosmia | 0.1 | 1.2 | 0.91 | 1.5 |
| Abdominal pain | 0.02 | 0.86 | 0.68 | 1.09 |

OR = odds ratio; CI= Confidence interval

## Discussion

The emergence of new SARS-CoV-2 mutants and recombinant strains unfortunately reduces the effectiveness of vaccines in the fight against the pandemic. For this reason, it is necessary to return to the beginning every 5-6 months in the fight against the epidemic. Although there is a vaccine, the number of patients who have had severe COVID-19 and have died is not small. Therefore, the importance of drug development and drug repositioning options is increasing in the fight against COVID-19. The development of effective treatment options against COVID-19 will only be possible with a clear understanding of the pathogenesis of COVID-19. Retinoic acid metabolism is defective in COVID-19 (cytokine storm), sepsis, ARDS and SIRS(10). Therefore, reconstitution of the signaling of retinoid may prove to be a valid strategy for COVID-19 management. Vitamin A is of special interest in the field of infectious diseases, especially for pulmonary infections(11). It is crucial for the development of normal lung tissue and tissue repair after injury due to infection [11]; therefore, it may play a role in recovery after severe COVID-19 pneumonia. Vitamin A has immune regulatory functions [12] and positively affects both the innate and adaptive immune cell response [13,14, 15]. Several studies revealed increased risks of severe illness due to respiratory tract infections in vitamin A-deficient individuals, whereas vitamin A supplementation can reduce the risks of severe illness and death, as was shown for children with measles and in influenza pneumonia in mice models [16–18]. The occurrence of severe infections and inflammation can also negatively affect vitamin A status, and several mechanisms such as urinary loss of vitamin A [19], decreased vitamin A hepatic mobilization [20], and reduced intestinal vitamin A absorption [21]. A recent study showed In hospitalized patients, the vitamin A plasma levels of critically ill patients were significantly reduced compared to patients with moderate disease (p < 0.05)(22). In patients with COVID-19, plasma levels of vitamin A decrease during acute inflammation, and severely reduced plasma levels of vitamin A are significantly associated with ARDS and mortality (35). Serum retinol levels were low in patients with severe COVID-19(36). Therefore, it is not surprising that ATRA exhibited antiviral effect against SARS-CoV-2 3CLpro cell culture (58) In addition to 13 CIS retinoic acid was found to protect against COVID-19 disease severity(59,60). 13 cis retinoic acid (isotretinoin), along with other retinoids, was found to inhibit SARS-CoV-2 replication in Vero E6 cells. According to Shoemark et al, 13 cis retinoic acid (isotretinoin) may bind to the SARS-CoV-2 spike protein at the SARS-CoV-2 spike fatty acid site, influencing the conformational changes required for receptor binding and stabilising the spike protein’s locked ACE2 conformation. In this way, isotretinoin may influence viral entry, especially in the early stages of infection (74).In our clinical study we selected 13 cis retinoic acid to be our candidate of investigation owing to its ability to inhibit Angiotensin II, IL-6, DHT, Platelet aggregation, thrombin production and ACE2. In addition to its ability to induce CD4 cells, IgA antibodies. 13-cis-retinoic acid is a member of the retinoid class of compounds which are structural analogues of vitamin A and include both natural and synthetic compounds. Naturally occurring retinoid compounds such as all trans retinoic acid (“ATRA”), 9-cis-retinoic acid, trans 3-4 didehydroretinoic acid, 4-oxo retinoic acid and retinol are pleiotrophic regulatory compounds that influence a large number of inflammatory, immune and structural cells(38). Here 13 cis retinoic acid was formulated as a powder aerosol, and its delivery to the lungs of COVID-19 patients was studied as well as oral 13 cis retinoic acid with the aim to explore its efficacy in treating COVID-19. This clinical study is the first clinical submitted on April 10, 2020, met QC Criteria on April 17, 2020, and was first published on April 20, 2020 (https://clinicaltrials.gov/ct2/keydates/NCT04353180).

According to our clinical findings, 13 cis retinoic acid, a synthetic vitamin A analogue, is effective in the treatment of COVID-19 with no mortality. All patients who received 13 cis retinoic acid noticed a high improvement (P<0.001), and the mean value for clinical improvement was 16.3±4.5 days. There was no significant difference regarding the laboratory parameters between before and after 14 days of treatment of patients had the standard of care treatment (P=0.66).

After treatment with Aerosolized and oral 13 cis retinoic acid there were significant decrease in Ang II level. Most of our patients were obese, conversely, adipose tissue is an important source of angiotensin, and obesity results in increased systemic RAS. ACE-Ang-II-AT1R axis, which has proinflammatory, profibrotic, prothrombotic, and vasoconstrictive effects, is potential mechanism of more severe SARS-CoV-2 infection(39). Plasma Ang II is associated with body weight, decreases during weight loss, and is associated with markers of insulin resistance in obese subjects with T2D(39). This agrees with our study, which revealed that obesity and diabetes were found to be risk factor for COVID-19 severity and mortality. Also, comes in agreement with many studies, which found that obesity is associated with severe disease and mortality in patients with coronavirus disease 2019 (COVID-19)(40,41,42,43). In the present study, TNF-a concentration was significantly higher in the 13 cis retinoic acid group in comparison to the other group after 14 days of treatment.this agrees with a recent study, which included 522 COVID patients and 40 healthy controls from two hospitals in Wuhan, China, which showed a negative correlation between T cells numbers and the serum IL6, IL-10, and TNF-a concentration, on the other hand, patients in decline period showing reduced IL-6, IL-10, and TNF-a concentrations and restored T cell counts (9).

Severe patterns of COVID-19 ARDS are characterized by thrombin burst and consequent coagulation activity(66).It was documented that retinoic acid significantly inhibit thrombin activity (65). This comes in agreement to our study, which revealed that thrombin activity significantly decreased in both groups, however, the reduction was significantly in the13 cis retinoic acid group than the other group.

The reduced vitamin A plasma levels were found to be associated with higher levels of inflammatory markers of acute SARS-COV-2 infection (CRP, ferritin). So, this comes in agreement to our study, which revealed that higher levels of inflammatory markers of acute SARS-COV-2 infection (CRP, ferritin) were significantly decreased after treatment in both groups, however, the reduction was significantly in the13 cis retinoic acid group than the other group.

Vitamin A deficient Mice has decreased numbers of IgA+ ASCs in site of the small bowel lamina propria. Consequently, mice deficient in retinoic acid(RA) precursor vitamin A lacked IgA antibodies-secreting cells in the small intestine(85,86). In addition, RA is considered to possess an activity of IgA isotype switching (87). Furthermore, retinoic acid, performing as a highly specific IgA isotype switch factor, cooperates with transforming growth factor beta 1 (TGFb1) to enhance the overall IgA response(88). Therefore, it is not surprising that isotretinoin (13-cis-retinoic acid) repair the function of IgA antibodies in submucosal pustular dermatosis patients(92). This agrees to our results, which showed that IgA significantly increased in both groups, however, the increase was significantly in the13 cis retinoic acid group than the other group.

Our study showed that decreased serum lymphocytes and the cholesterol levels of COVID-19, which responded to the treatment and increased significantly in both groups, however, the increase was significantly in the13 cis retinoic acid group than the other group.

Also, it was founded that decreased serum lymphocytes responded to the treatment and increased(134).Also, Researchers from Wenzhou, China looked at clinical laboratory investigations in patients with COVID-19. They showed a marked reduction in the cholesterol levels of COVID-19(93). Also, during the early stages of infection, it was found that cholesterol levels decline rapidly and increase as the patient starts to recover. Intracellular cholesterol level is regulated by two competing pathways, cholesterol uptake and efflux, and ATP-binding cassette transporter (ABCA1) plays a major role in the cholesterol efflux pathways. Retinoic Acid induces macrophage cholesterol efflux and inhibits atherosclerotic plaque formation in apoE-deficient mice, was reported in a study(95). All trans-retinoic acid (ATRA) and 13-cis-RA may play an important role in immune response(97).

Increased ANG II levels in COVID-19 has been reported (44). A study by Liu and colleagues (2020), observed markedly high ANG II levels in COVID-19 patients when compared to healthy controls. Many studies found that plasma ACE 2 increased in COVID-19 patients (48,49,50). In our study plasma ACE2 was significantly decreased after treatment in both groups, but, it was significantly reduced after treatment in the13 cis retinoic acid group than the other group.

Dihydrotestosterone (DHT) was found to be a potent activator of TMPRSS2(118) and this activation may be followed by processing and activation of COVID-19 spike protein to bind to its ACE2 receptors in the lung, kidney, other organs. A study demonstrated that 13-cis-retinoic acid competitively and reversibly inhibits dihydrotestosterone (124). Also, in the current study Dihydrotestosterone (DHT) was significantly decreased after treatment in the13 cis retinoic acid group only, also, it was significantly reduced after treatment in the13 cis retinoic acid group than the other group. AngII via AT1 receptors upregulates many proinflammatory genes, such as vascular cell adhesion molecule-1 (VCAM-1), intercellular adhesion molecule-1 (ICAM-1), interleukin-6 (IL6) (54), but 13-cis-RA specifically downregulated the AT1 protein in a dose- and time-dependent manner. Deregulation of the AT1 expression results in reducing AngII-mediated intracellular calcium release. Similarly with receptor downregulation, treatment with 13cRA resulted in a significant reduction in AT1 mRNA (55).

In our study soluble ACE2 was found to be decreased after treatment with 13 cis retinoic acid. This is similar to a study, which investigated the effects of the 672 clinically approved drugs in CMAP found that (13cRA) is the strongest down-regulator of ACE2 suggesting its therapeutic potential in preventing the entry of COVID-2019 to the host cell (56).

Serum interleukin-6 is one of the most important factor implicated in cytokine storm and is an indicator for severity in patients with SARS-CoV-2 infection(57). Our results agreed many results in which 13 cis retinoic acid (Isotretinoin) therapy significantly decreased the secretion of IL-1b, IL-6, IL-10, and IL-12p70 by monocytes from acne patients in response to P. acnes compared with pretreatment levels. Cytokine secretion decreased as early as 1 week after 13 cis retinoic acid(isotretinoin) therapy and continued to decrease through 20 weeks of therapy (58). A recent study reported that retinoic acid have preventive effects on pulmonary fibrosis by inhibiting IL-6-dependent proliferation and TGF-b1-dependent trans differentiation of lung fibroblasts (59). Also, another study reported that 13-cisretinoic acid and other retinoid analogs inhibit IL-1-induced IL6 production and that this effect is analog-specific and, at least partially, transcriptionally mediated (60). This is agreement to our study, which revealed that IL-6 were significantly reduced after the treatment. The previous results, explains our results, which revealed that, mortality and need of ICU were significantly lower and also, the mean duration to clinical improvement and hospital discharge were lower in the13 cis retinoic acid.

Lastly, we conclude that 13 cis retinoic acid, will be a promising drug in the treatment of COVID19.

## Data Availability

All data produced in the present work are contained in the manuscript

## Limitation of the study

The small number of patients and the fact that the study was conducted in a single center. Therefore, we need multicenter studies on a large patient scale. Due to the potential teratogenic effect of 13-Cis retinoic acid, pregnant female patients were not included in the study. RAs are the most potent teratogenic agents. RA is not given to a patient who is planning pregnancy regardless of the dose, even 6 months before. and pregnancy is prevented for 6 months, sometimes up to a year, after the treatment is stopped. The reason for this is the understanding that RAs are stored in adipose tissue. Pregnancy allowed after RA’s are cleared from the bod**y**

## Disclosure

The authors report no conflict of interest in this work.

## Notes

### Competing Interest Statement

The authors have declared no competing interest.

### Clinical Trial

NCT04353180

### Funding Statement

This study did not receive any funding

### Author Declarations

The Research Ethics Committee of the Faculty of Medicine, Kafrelsheikh University, EGYPT.

## References

1 Zhu N, Zhang DY, Wang WL, et al. A novel coronavirus from patients with pneumonia in China, 2019. N Engl J Med (2020). doi: 10.1056/nejmoa2001017

2 Huang C, Wang Y, Li X, Ren L, Zhao J, Hu Y, et al. Clinical features of patients infected with 2019 novel coronavirus in Wuhan, China. Lancet (2020) 395(10223):497–506. doi: 10.1016/S0140-6736(20)30183-5

3 Lai CC, Shih TP, Ko WC, Tang HJ, Hsueh PR. Severe acute respiratory syndrome coronavirus 2 (SARS-CoV-2) and coronavirus disease-2019 (COVID-19): The epidemic and the challenges. Int J Antimicrob Agents (2020) 55(3):105924. doi: 10.1016/j.ijantimicag.2020.105924

4 Ni W, Yang X, Yang D, et al. Role of angiotensin-converting enzyme 2 (ACE2) in COVID-19. Crit Care (2020) 24:422. doi: 10.1186/s13054-020-03120-0

5 Hoffmann M, Kleine-Weber H, Schroeder S, Krüger N, Herrler T, Erichsen S, et al. SARS-CoV-2 Cell Entry Depends on ACE2 and TMPRSS2 and Is Blocked by a Clinically Proven Protease Inhibitor. Cell (2020) 181(2):271– 280.e8. doi: 10.1016/j.cell.2020.02.052

6 Li CK, Wu H, Yan H, et al. T cell responses to whole SARS coronavirus in humans. J Immunol (2008) 181(8):5490–500. doi: 10.4049/jimmunol.181.8.5490

7 Li G, Fan Y, Lai Y, Han T. First Coronavirus infections and immune responses. (2020). doi: 10.1002/jmv.25685

8 Mubarak A, Alturaiki W, Hemida MG. Middle East Respiratory Syndrome Coronavirus (MERS-CoV): Infection, Immunological Response, and Vaccine Development. J Immunol Res (2019) 2019:6491738. doi: 10.1155/2019/6491738

9 Diao B, Wang C, Tan Y, Chen X, Liu Y, Ning L, et al. Reduction and Functional Exhaustion of T Cells in Patients With Coronavirus Disease 2019 (COVID-19). Front Immunol (2020) 11:827:827. doi: 10.3389/fimmu.2020.00827

10 Mahmoud Elkazzaz, Tamer Haydara, Yousry Esam-Eldin Abo-Amer et al. STRA6 (vitamin A receptor), as a Novel binding receptor of COVID-19 (A breakthrough), 13 September 2021, PREPRINT (Version 3) available at Research Square [https://doi.org/10.21203/rs.3.rs-892203/v3]

11 Review Vitamin A Deficiency and the Lung. Timoneda J, Rodríguez-Fernández L, Zaragozá R, Marín MP, Cabezuelo MT, Torres L, Viña JR, Barber TNutrients. 2018 Aug 21; 10(9):.

12 Review Carotenoid action on the immune response. Chew BP, Park J J Nutr. 2004 Jan; 134(1):257S–261S.

13 Review Role of Vitamin A in the Immune System. Huang Z, Liu Y, Qi G, Brand D, Zheng SG J Clin Med. 2018 Sep 6; 7(9):.

14 Review Modulation of T cell and innate immune responses by retinoic Acid.Raverdeau M, Mills KHJ Immunol. 2014 Apr 1; 192(7):2953–8.

15 Vitamin A deficiency disrupts vaccine-induced antibody-forming cells and the balance of IgA/IgG isotypes in the upper and lower respiratory tract. Surman SL, Rudraraju R, Sealy R, Jones B, Hurwitz JL Viral Immunol. 2012 Aug; 25(4):341–4.

16 Vitamin A supplementation in infectious diseases: a meta-analysis. Glasziou PP, Mackerras DE BMJ. 1993 Feb 6; 306(6874):366–70.

17 17-A randomized, controlled trial of vitamin A in children with severe measles.Hussey GD, Klein MN Engl J Med. 1990 Jul 19; 323(3):160–4.

18 Effect of Vitamin A Deficiency in Dysregulating Immune Responses to Influenza Virus and Increasing Mortality Rates After Bacterial Coinfections. Penkert RR, Smith AP, Hrincius ER, McCullers JA, Vogel P, Smith AM, Hurwitz JLJ Infect Dis. 2021 May 28; 223(10):1806–1816.

19 Review Vitamin A, infection, and immune function. Stephensen CBAnnu Rev Nutr. 2001; 21():167–92.

20 Model-based compartmental analysis indicates a reduced mobilization of hepatic vitamin A during inflammation in rats. Gieng SH, Green MH, Green JB, Rosales FJJ Lipid Res. 2007 Apr; 48(4):904–13.

21 Model-based compartmental analysis indicates a reduced mobilization of hepatic vitamin A during inflammation in rats.Gieng SH, Green MH, Green JB, Rosales FJJ Lipid Res. 2007 Apr; 48(4):904–13

22 Tepasse, P. R., Vollenberg, R., Fobker, M., Kabar, I., Schmidt, H., Meier, J. A., Nowacki, T., & Hüsing-Kabar, A. (2021). Vitamin A Plasma Levels in COVID-19 Patients: A Prospective Multicenter Study and Hypothesis. Nutrients, 13(7), 2173. https://doi.org/10.3390/nu13072173

23 Mahmoud Elkazzaz, Tamer Haydara, Yousry Esam-Eldin Abo-Amer et al. STRA6, as A Novel Binding Receptor of COVID-19, A Breakthrough That could Explain COVID-19 Symptoms with Unknown Aetiology., 27 September 2021, PREPRINT (Version 1) available at Research Square [https://doi.org/10.21203/rs.3.rs-936697/v1]

24 Chen, Y., Clarke, O. B., Kim, J., Stowe, S., Kim, Y. K., Assur, Z., Cavalier, M., Godoy-Ruiz, R., von Alpen, D. C., Manzini, C., Blaner, W. S., Frank, J., Quadro, L., Weber, D. J., Shapiro, L., Hendrickson, W. A., & Mancia, F. (2016). Structure of the STRA6 receptor for retinol uptake. Science (New York, N.Y.), 353(6302), aad8266. https://doi.org/10.1126/science.aad8266.

25 Kelly, Mary, and Johannes von Lintig. “STRA6: role in cellular retinol uptake and efflux.” Hepatobiliary surgery and nutrition vol. 4,4 (2015): 229–42. doi:10.3978/j.issn.2304-3881.2015.01.12

26 Barrett, Crisha et al. “Human Peripheral Blood Mononuclear Cells Express High Levels of the Vitamin a Transport Protein, Stimulated by Retinoic Acid 6 (P19-004-19).” Current Developments in Nutrition vol. 3,Suppl 1 nzz049.P19–004-19. 13 Jun. 2019, doi:10.1093/cdn/nzz049.P19-004-19

27 Barrett, Crisha et al. “Human Peripheral Blood Mononuclear Cells Express High Levels of the Vitamin a Transport Protein, Stimulated by Retinoic Acid 6 (P19-004-19).” Current Developments in Nutrition vol. 3,Suppl 1 nzz049.P19–004-19. 13 Jun. 2019, doi:10.1093/cdn/nzz049.P19-004-19

28 Kelly, M., Widjaja-Adhi, M. A., Palczewski, G., & von Lintig, J. (2016). Transport of vitamin A across blood-tissue barriers is facilitated by STRA6. FASEB journal : official publication of the Federation of American Societies for Experimental Biology, 30(8), 2985–2995. https://doi.org/10.1096/fj.201600446R

29 STRA6 is critical for cellular vitamin A uptake and homeostasis. Amengual J, Zhang N, Kemerer M, Maeda T, Palczewski K, Von Lintig J Hum Mol Genet. 2014 Oct 15; 23(20):5402–17.

30 The STRA6 receptor is essential for retinol-binding protein-induced insulin resistance but not for maintaining vitamin A homeostasis in tissues other than the eye.Berry DC, Jacobs H, Marwarha G, Gely-Pernot A, O’Byrne SM, DeSantis D, Klopfenstein M, Feret B, Dennefeld C, Blaner WS, Croniger CM, Mark M, Noy N, Ghyselinck NB J Biol Chem. 2013 Aug 23; 288(34):24528–39.

31 Developmental expression pattern of Stra6, a retinoic acid-responsive gene encoding a new type of membrane protein. Bouillet P, Sapin V, Chazaud C, Messaddeq N, Décimo D, Dollé P, Chambon P Mech Dev. 1997 May; 63(2):173–86.

32 Receptor-mediated cellular uptake mechanism that couples to intracellular storage. Kawaguchi R, Yu J, TerStepanian M, Zhong M, Cheng G, Yuan Q, Jin M, Travis GH, Ong D, Sun HACS Chem Biol. 2011 Oct 21; 6(10):1041–51.

33 RBP4 disrupts vitamin A uptake homeostasis in a STRA6-deficient animal model for Matthew-Wood syndrome. Isken A, Golczak M, Oberhauser V, Hunzelmann S, Driever W, Imanishi Y, Palczewski K, von Lintig J Cell Metab. 2008 Mar; 7(3):258–68.

34 Case-control analysis of SNPs in GLUT4, RBP4 and STRA6: association of SNPs in STRA6 with type 2 diabetes in a South Indian population. Nair AK, Sugunan D, Kumar H, Anilkumar G PLoS One. 2010 Jul 6; 5(7):e11444. Page 22/32

35 Pasutto, F., Sticht, H., Hammersen, G., Gillessen-Kaesbach, G., Fitzpatrick, D. R., Nürnberg, G., Brasch, F., SchirmerZimmermann, H., Tolmie, J. L., Chitayat, D., Houge, G., Fernández-Martínez, L., Keating, S., Mortier, G., Hennekam, R. C. M., von der Wense, A., Slavotinek, A., Meinecke, P., Bitoun, P., … Rauch, A. (2007). Mutations in STRA6 cause a broad spectrum of malformations including anophthalmia, congenital heart defects, diaphragmatic hernia, alveolar capillary dysplasia, lung hypoplasia, and mental retardation. American Journal of Human Genetics, 80(3), 550-60. https://doi.org/10.1086/512203

36 Tepasse PR, Vollenberg R, Fobker M, Kabar I, Schmidt H, Meier JA, Nowacki T, Hüsing-Kabar A. Vitamin A Plasma Levels in COVID-19 Patients: A Prospective Multicenter Study and Hypothesis. Nutrients. 2021 Jun 24;13(7):2173. doi: 10.3390/nu13072173; PMID: 34202697; PMCID: PMC8308355.

37 izRodan Sarohan, Hakan Akelma, Eşref Araç, Özgür AslanmedRxiv 202 1.01.30.21250844; doi: https://doi.org/10.1101/2021.01.30.21250844

38 Bushue, N., & Wan, Y. J. (2010). Retinoid pathway and cancer therapeutics. Advanced drug delivery reviews, 62(13), 1285–1298. https://doi.org/10.1016/j.addr.2010.07.003

39 Saiki A, Ohira M, Endo K, Koide N, Oyama T, Murano T, Watanabe H, Miyashita Y, Shirai K. Circulating angiotensin II is associated with body fat accumulation and insulin resistance in obese subjects with type 2 diabetes mellitus. Metabolism. 2009 May;58(5):708–13. doi: 10.1016/j.metabol.2009.01.013; PMID: 19375596.

40 Cai, Z., Yang, Y. & Zhang, J. Obesity is associated with severe disease and mortality in patients with coronavirus disease 2019 (COVID-19): a meta-analysis. BMC Public Health 21, 1505 (2021). https://doi.org/10.1186/s12889-021-11546-6

41 Zhao X, Gang X, He G, Li Z, Lv Y, Han Q, Wang G. Obesity Increases the Severity and Mortality of Influenza and COVID-19: A Systematic Review and Meta-Analysis. Front Endocrinol (Lausanne). 2020 Dec 21;11:595109. doi: 10.3389/fendo.2020.595109; PMID: 33408692; PMCID: PMC7779975.

42 Zhang, X., Lewis, A.M., Moley, J.R. et al. A systematic review and meta-analysis of obesity and COVID-19 outcomes. Sci Rep 11, 7193 (2021). https://doi.org/10.1038/s41598-021-86694-1

43 Poly TN, Islam MM, Yang HC, Lin MC, Jian WS, Hsu MH, Jack Li YC. Obesity and Mortality Among Patients Diagnosed With COVID-19: A Systematic Review and Meta-Analysis. Front Med (Lausanne). 2021 Feb 5;8:620044. doi: 10.3389/fmed.2021.620044; PMID: 33634150; PMCID: PMC7901910.

44 Aksoy H, Karadag AS, Wollina U. Angiotensin II receptors: Impact for COVID-19 severity. Dermatol Ther. 2020 Nov;33(6):e13989. doi: 10.1111/dth.13989. Epub 2020 Jul 27; PMID: 32645228; PMCID: PMC7361069.

45 Clinical and biochemical indexes from 2019-nCoV infected patients linked to viral loads and lung injury.Liu Y, Yang Y, Zhang C, Huang F, Wang F, Yuan J, Wang Z, Li J, Li J, Feng C, Zhang Z, Wang L, Peng L, Chen L, Qin Y, Zhao D, Tan S, Yin L, Xu J, Zhou C, Jiang C, Liu L Sci China Life Sci. 2020 Mar; 63(3):364–374.

46 The Serum Profile of Hypercytokinemia Factors Identified in H7N9-Infected Patients can Predict Fatal Outcomes. Guo J, Huang F, Liu J, Chen Y, Wang W, Cao B, Zou Z, Liu S, Pan J, Bao C, Zeng M, Xiao H, Gao H, Yang S, Zhao Y, Liu Q, Zhou H, Zhu J, Liu X, Liang W, Yang Y, Zheng S, Yang J, Diao H, Su K, Shao L, Cao H, Wu Y, Zhao M, Tan S, Li H, Xu X, Wang C, Zhang J, Wang L, Wang J, Xu J, Li D, Zhong N, Cao X, Gao GF, Li L, Jiang C.Sci Rep. 2015 Jun 1; 5():10942.

47 Clinical features of patients infected with 2019 novel coronavirus in Wuhan, China.Huang C, Wang Y, Li X, Ren L, Zhao J, Hu Y, Zhang L, Fan G, Xu J, Gu X, Cheng Z, Yu T, Xia J, Wei Y, Wu W, Xie X, Yin W, Li H, Liu M, Xiao Y, Gao H, Guo L, Xie J, Wang G, Jiang R, Gao Z, Jin Q, Wang J, Cao B.Lancet. 2020 Feb 15; 395(10223):497–506

48 Patel SK, Juno JA, Lee WS, Wragg KM, Hogarth PM, Kent SJ, et al. Plasma ACE2 activity is persistently elevated following SARS-CoV-2 infection: implications for COVID-19 pathogenesis and consequences. European Respiratory Journal. 2021:2003730. pmid:33479113

49 Lundström A, Ziegler L, Havervall S, Rudberg A-S, von Meijenfeldt F, Lisman T, et al. Soluble angiotensin-converting enzyme 2 is transiently elevated in COVID-19 and correlates with specific inflammatory and endothelial markers. medRxiv.

50 Nagy B Jr., Fejes Z, Szentkereszty Z, Suto R, Varkonyi I, Ajzner E, et al. A dramatic rise in serum ACE2 activity in a critically ill COVID-19 patient. Int J Infect Dis. 2021;103:412–4. pmid:33249290

51 Xu H, Zhong L, Deng J, et al. High expression of ACE2 receptor of 2019-nCoV on the epithelial cells of oral mucosa. Int J Oral Sci (2020) 12(1):8. doi: 10.1038/s41368-020-0074-x

52 Zhao Y, Zhao Z, Wang Y, Zhou Y, Ma Y, Zuo W. Single-Cell RNA Expression Profiling of ACE2, the Receptor of SARS-CoV-2. Am J Respir Crit Care Med (2020) 202(5):756–9. doi: 10.1164/rccm.202001-0179LE

53 Straus MR, Tang T, Lai AL, Flegel A, Bidon M, Freed JH, et al. Ca2+ Ions Promote Fusion of Middle East Respiratory Syndrome Coronavirus with Host Cells and Increase Infectivity. J Virol (2020) 94(13):e00426–20. doi: 10.1128/JVI.00426-20

54 Nitschke R, Henger A, Ricken S, Gloy J, Müller V, Greger R, et al. Angiotensin II increases the intracellular calcium activity in podocytes of the intact glomerulus. Kidney Int (2000) 57(1):41–9. doi: 10.1046/j.1523-1755.2000.00810.x

55 R Thekkumkara T. 13-cis-Retinoic acid specific down-regulation of angiotensin type 1 receptor in rat liver epithelial and aortic smooth muscle cells. J Mol Endocrinol (2012) 48(2):99–114. doi: 10.1530/JME-11-0095

56 Sinha, Sanju et al. “In vitro and in vivo identification of clinically approved drugs that modify ACE2 expression.” Molecular systems biology vol. 16,7 (2020): e9628. doi:10.15252/msb.20209628

57 Zhang, J., Hao, Y., Ou, W. et al. Serum interleukin-6 is an indicator for severity in 901 patients with SARS-CoV-2 infection: a cohort study. J Transl Med 18, 406 (2020). https://doi.org/10.1186/s12967-020-02571-x

58 Dispenza MC, Wolpert EB, Gilliland KL, Dai JP, Cong Z, Nelson AM, Thiboutot DM. Systemic isotretinoin therapy normalizes exaggerated TLR-2-mediated innate immune responses in acne patients. J Invest Dermatol. 2012 Sep;132(9):2198–205. doi: 10.1038/jid.2012.111. Epub 2012 Apr 19; PMID: 22513780; PMCID: PMC3614089.

59 Tabata C, Tabata R, Nakano T. The calpain inhibitor calpeptin prevents bleomycin-induced pulmonary fibrosis in mice. Clin Exp Immunol (2010) 162(3):560– 567. doi: 10.1111/j.13652249.2010.04257

60 Zitnik RJ, Kotloff RM, Latifpour J, Zheng T, Whiting NL, Schwalb J, et al. J Immunol (1994) 152(3):1419–27.

61 Skariyachan S, Challapilli SB, Packirisamy S, Kumargowda ST, Sridhar VS. Front Microbiol (2019). doi: 10.3389/fmicb.2019.00569

62 Totura AL, Whitmore A, Agnihothram S, Schäfer A, Katze MG, Heise MT, et al. mBio (2015) 6(3):e00638.–15. doi: 10.1128/mBio.00638-15

63 Liang Z, Wu S, Li Y, et al. Activation of Toll-like receptor 3 impairs the dengue virus serotype 2 replication through induction of IFN-b in cultured hepatoma cells. PloS One (2011) 6(8):e23346. doi: 10.1371/journal.pone.0023346

64 Chuang HC, Lin HY, Liao PL, Huang CC, Lin LL, Hsu WM, et al. Immunomodulator polyinosinic-polycytidylic acid enhances the inhibitory effect of 13-cis-retinoic acid on neuroblastoma through a TLR3-related immunogenic-apoptotic response. Lab Invest (2020) 100(4):606–18. doi: 10.1038/s41374-019-0356-0

65 Anantha Krishna, T. H., Kamalraj, S., Anikisetty, M., Naidu, K. A., Surin, W. R., & Jayabaskaran, C. (2019). Inhibition of thrombin, an unexplored function of retinoic acid. Biochemistry and biophysics reports, 18, 100636. https://doi.org/10.1016/j.bbrep.2019.100636

66 Ranucci, M., Sitzia, C., Baryshnikova, E., Di Dedda, U., Cardani, R., Martelli, F., & Corsi Romanelli, M. (2020). Covid-19-Associated Coagulopathy: Biomarkers of Thrombin Generation and Fibrinolysis Leading the Outcome. Journal of clinical medicine, 9(11), 3487. https://doi.org/10.3390/jcm9113487.

67 Levi M, Thachil J, Iba T, Levy JH. Coagulation abnormalities and thrombosis in patients with COVID-19. Lancet Haematol (2020) 7(6):e438– 40. doi: 10.1016/S2352-3026(20)30145-9

68 Zhou F, et al. Clinical course and risk factors for mortality of adult inpatients with COVID-19 in Wuhan, China: a retrospective cohort study. Lancet (2020) 395:1054–62. doi: 10.1016/S0140-6736(20)30566-3

69 Borissoff JI, Spronk HM, Heeneman S, ten Cate H. Is thrombin a key player in the ‘coagulation-atherogenesis’ maze? Cardiovasc Res (2009) 82(3):392– 403.doi: 10.1093/cvr/cvp066

71 Anantha Krishna TH, Kamalraj S, Anikisetty M, Naidu KA, Surin WR, Jayabaskaran C. Inhibition of thrombin, an unexplored function of retinoic acid. Biochem Biophys Rep (2019) 18:100636. doi: 10.1016/j.bbrep.2019.100636

72 Camacho M, Rodriguez C, Salazar J, Mart inez-Gonza lez J, Ribalta J, Escudero JR, et al. Retinoic acid induces PGI synthase expression in human endothelial cells. J Lipid Res (2008) 49(8):1707–14. doi: 10.1194/jlr.M700559-JLR200

73 Oh DK, Kim YS, Oh YM. Lung Regeneration Therapy for Chronic Obstructive Pulmonary Disease. Tuberc Respir Dis (Seoul) (2017) 80(1):1– 10. doi: 10.4046/trd.2017.80.1.1

74 Shoemark DK, Colenso CK, Toelzer C, et al. Molecular Simulations suggest Vitamins, Retinoids and Steroids as Ligands of the Free Fatty Acid Pocket of the SARS-CoV-2 Spike Protein*. Angew Chem Int Ed Engl. 2021;60(13):7098–7110. https://doi.org/10.1002/anie.202015639

75 Chao YX, Rötzschke O, Tan EK. The role of IgA in COVID-19. Brain Behav Immun (2020) 87:182–3. doi: 10.1016/j.bbi.2020.05.057

76 Petherick A. Developing antibody tests for SARS-CoV-2. Lancet (2020) 395 (10230):1101–2. doi: 10.1016/S0140-6736(20)30788-1

76 Kumar N, Arthur CP, Ciferri C, Matsumoto ML. Structure of the secretory immunoglobulin A core. Science (2020) 367(6481):1008–14. doi: 10.1126/science.aaz5807

77 Iwata M, Hirakiyama A, Eshima Y, Kagechika H, Kato C, Song SY. Retinoic acid imprints gut-homing specificity on T cells. Immunity (2004) 21(4):527– 38. doi: 10.1016/j.immuni.2004.08.011

78 Mora JR, Iwata M, Eksteen B, Song u T, Senman B, et al. Generation of gut-homing IgA-secreting B cells by intestinal dendritic cells. Science (2006) 314(5802):1157–60. doi: 10.1126/science.1132742

79 Tokuyama H, Tokuyama Y. The regulatory effects of all-trans-retinoic acid on isotype switching: retinoic acid induces IgA switch rearrangement in cooperation with IL-5 and inhibits IgG1 switching. Cell Immunol (1999) 192:41–7. doi: 10.1006/cimm.1998.1438

80 Wiedermann U, Hanson LA, Kahu H, Dahlgren UI. Aberrant T-cell function in vitro and impaired T-cell dependent antibody response in vivo in vitamin A-deficient rats. Immunology (1993) 80(4):581–6.

81 Sirisinha S, Darip MD, Moongkarndi P, Ongsakul M, Lamb AJ. Impaired local immune response in vitamin A-deficient rats. Clin Exp Immunol (1980) 40(1):127–35.

82 Stephan U, König W. Effect of antigens from Nippostrongylus brasiliensis and cytokines on the ongoing IgE synthesis in vitro. Int Arch Allergy Immunol (1992) 98(4):299–307. doi: 10.1159/000236202

83 Dauwerse JG, Jumelet EA, Wessels JW, Saris JJ, Hagemeijer A, Beverstock GC, et al. Extensive cross-homology between the long and the short arm of chromosome 16 may explain leukemic inversions and translocations. Blood (1992) 79(5):1299–304.

84 Mora JR, von Andrian UH. Differentiation and homing of IgA-secreting cells. Mucosal Immunol (2008) 1(2):96–109. doi: 10.1038/mi.2007.14

85 Nikawa T, Odahara K, Koizumi H, Kido Y, Teshima S, Rokutan K, et al. Vitamin A prevents the decline in immunoglobulin A and Th2 cytokine levels in small intestinal mucosa of protein-malnourished mice. J Nutr (1999) 129(5):934–41. doi: 10.1093/jn/129.5.934

86 Sato A, Hashiguchi M, Toda E, Iwasaki A, Hachimura S, Kaminogawa S. CD11b+ Peyer’s patch dendritic cells secrete IL-6 and induce IgA secretion from naive B cells. J Immunol (2003) 171(7):3684–90. doi: 10.4049/jimmunol.171.7.3684

87 Park MH, Park SR, Lee MR, et al. Retinoic acid induces expression of Ig germ line a transcript, an IgA isotype switching indicative, through retinoic acid receptor. Genes Genom (2011) 33:83–8. doi: 10.1007/s13258-010-0168-5

88 Seo GY, Jang YS, Kim HA, Lee MR, Park MH, Park SR, et al. Retinoic acid, acting as a highly specific IgA isotype switch factor, cooperates with TGF-b1 to enhance the overall IgA response. J Leukoc Biol (2013) 94(2):325–35. doi: 10.1189/jlb.0313128

89 Lee JM, Jang YS, Jin BR, Kim SJ, Kim HJ, Kwon BE, et al. Retinoic acid enhances lactoferrin-induced IgA responses by increasing betaglycan expression. Cell Mol Immunol (2016) 13(6):862–70. doi: 10.1038/cmi.2015.73

90 Villamor E, Fawzi WW. Effects of vitamin a supplementation on immune responses and correlation with clinical outcomes. Clin Microbiol Rev (2005) 18(3):446–64. doi: 10.1128/CMR.18.3.446-464.2005

91 Semba RD, Ndugwa C, Perry RT, Clark TD, Jackson JB, Melikian G, et al. Effect of periodic vitamin A supplementation on mortality and morbidity of human immunodeficiency virus-infected children in Uganda: A controlled clinical trial. Nutrition (2005) 21(1):25–31. doi: 10.1016/j.nut.2004.10.004

92 Gruss C, Zillikens D, Hashimoto T, Amagai M, Kroiss M, Vogt T, et al. Rapid response of IgA pemphigus of subcorneal pustular dermatosis type to treatment with isotretinoin. J Am Acad Dermatol (2000) 43(5 Pt 2):923–6. doi: 10.1067/mjd.2000.104002

93 Xingzhong H, Dong C, Lianpeng W, Guiqing H, Wei Y. Low Serum Cholesterol Level Among Patients with COVID-19 Infection in Wenzhou, China (February 21, 2020). Available at: https://ssrn.com/abstract=3544826. doi: 10.2139/ssrn.3544826

94 Ravnskov U. High cholesterol may protect against infections and atherosclerosis. QJM: Int J Med (2003) 96(12):927–34. doi: 10.1093/qjmed/hcg150

95 Zhou W, Lin J, Chen H, Wang J, Liu Y, Xia M. Retinoic acid induces macrophage cholesterol efflux and inhibits atherosclerotic plaque formation in apoE-deficient mice. Br J Nutr (2015) 114(4):509–18. doi: 10.1017/S0007114515002159

96 Jiang H, Badralmaa Y, Yang J, Lempicki R, Hazen A, Natarajan V. Retinoic acid and liver X receptor agonist synergistically inhibit HIV infection in CD4 + T cells by up-regulating ABCA1-mediated cholesterol efflux. Lipids Health Dis (2012) 11:69. doi: 10.1186/1476-511X-11-69

97 Oliveira LM, Teixeira FME, Sato MN. Impact of Retinoic Acid on Immune Cells and Inflammatory Diseases. Mediators Inflamm (2018) 2018:3067126. doi: 10.1155/2018/3067126

98 Harms M, Péchère M, Krischer J, Studer E, Saurat JH. Oral isotretinoin in HIV-positive women with acne: report of three cases. Dermatology (1998) 196(1):163–4. doi: 10.1159/000017853

99 Scully EP, Haverfield J, Ursin RL, Tannenbaum C, Klein SL. Considering how biological sex impacts immune responses and COVID-19 outcomes. Nat Rev Immunol (2020) 20(7):442–7. doi: 10.1038/s41577-020-0348-8

100 Meng Y, Wu P, Lu W, Liu K, Ma K, Huang L, et al. Sex-specific clinical characteristics and prognosis of coronavirus disease-19 infection in Wuhan, China: A retrospective study of 168 severe patients. PloS Pathog (2020) 16 (4):e1008520. doi: 10.1371/journal.ppat.1008520

101 Takahashi T, Wong P, Ellingson MK, Lucas C, Klein J, Israelow B, et al. Yale IMPACT research team. Sex differences in immune responses to SARSCoV-2 that underlie disease outcomes. medRxiv (2020) 9. doi: 10.1101/2020.06.06.201234142020.06.06.20123414.

102 Recchia F, Saggio G, Cesta A, Candeloro G, Nuzzo A, Lombardo M, et al. Interleukin-2 and 13-cis retinoic acid as maintenance therapy in advanced ovarian cancer. Int J Oncol (2005) 27(4):1039–46. doi: 10.3892/ijo.27.4.1039

103 Recchia F, Saggio G, Cesta A, et al. Phase II study of interleukin-2 and 13-cisretinoic acid as maintenance therapy in metastatic colorectal cancer. Cancer Immunol Immunother (2007) 56:699–708. doi: 10.1007/s00262-006-0224-9

104 Pomponi F, Cariati R, Zancai P, De Paoli P, Rizzo S, Tedeschi RM, et al. Retinoids irreversibly inhibit in vitro growth of Epstein-Barr virusimmortalized B lymphocytes. Blood (1996) 88(8):3147–59. doi: 10.1182/blood.V88.8.3147.bloodjournal8883147

105 Hammerschmidt SI, Friedrichsen M, Boelter J, Lyszkiewicz M, Kremmer E, Pabst O, et al. Retinoic acid induces homing of protective T and B cells to the gut after subcutaneous immunization in mice. J Clin Invest (2011) 121 (8):3051–61. doi: 10.1172/JCI44262

106 Laforge M, Elbim C, Frère C, Hémadi M, Massaad C, Nuss P, et al. Tissue damage from neutrophil-induced oxidative stress in COVID-19. Nat Rev Immunol (2020) 20(9):515–6. doi: 10.1038/s41577-020-0407-1

107 Veras FP, Pontelli MC, Silva CM, Toller-Kawahisa JE, de Lima M, Nascimento DC, et al. SARS-CoV-2-triggered neutrophil extracellular traps mediate COVID-19 pathology. J Exp Med (2020) 217(12):e20201129. doi: 10.1084/jem.20201129

108 Norris DA, Osborn R, Robinson W, Tonnesen MG. Isotretinoin produces significant inhibition of monocyte and neutrophil chemotaxis in vivo in patients with cystic acne. J Invest Dermatol (1987) 89(1):38–43. doi: 10.1111/1523-1747.ep12580370

109 Dey D, Borkotoky S, Banerjee M. In silico identification of Tretinoin as a SARS-CoV-2 envelope (E) protein ion channel inhibitor. Comput Biol Med. 2020 Dec;127:104063. doi: 10.1016/j.compbiomed.2020.104063. Epub 2020 Oct 20; PMID: 33126128; PMCID: PMC7574788.

110 Patrì A, Fabbrocini G. Possible role of neuropilins in dermatological manifestations of SARS-CoV-2 infection. Int J Dermatol. 2021. https://doi.org/10.1111/ijd.15546

111 Bauer W, Diehl-Wiesenecker E, Ulke J, et al. Outcome prediction by serum calprotectin in patients with COVID-19 in the emergency department. J Infect. 2021;82(4):84–123. https://doi.org/10.1016/j.jinf.2020.11.016

112 Gundogdu M, Dere G. Is systemic isotretinoin use a risk factor for coronavirus disease 2019 (COVID-19)? J Cosmet Dermatol. 2021;20(6):1568–1570. https://doi.org/10.1111/jocd.14044

113 Donnarumma M, Nocerino M, Lauro W, Annunziata MC, Marasca C, Fabbrocini G. Isotretinoin in acne treatment during the coronavirus disease 2019 (COVID-19): A retrospective analysis of adherence to therapy and side effects. Dermatol Ther. 2021;34(1):e14677. https://doi.org/10.1111/dth.14677

114 Buckley JL, Chastain MA, Rietschel RL. Improvement of cystic fibrosis during treatment with isotretinoin. SKINmed. 2006;5(5):252–225. https://doi.org/10.1111/j.1540-9740.2006.04514.x

115 Fauchère JC, Ersch J, Grant DA, Zimmermann R, Bucher HU, Stallmach T. Acceleration of lung maturation in a human fetus following maternal isotretinoin intake. Biol Neonate. 2006;90(3):203–206. https://doi.org/10.1159/000093589

116 Leach, D.A., Mohr, A., Giotis, E.S. et al. The antiandrogen enzalutamide downregulates TMPRSS2 and reduces cellular entry of SARS-CoV-2 in human lung cells. Nat Commun 12, 4068 (2021). https://doi.org/10.1038/s41467-021-24342-y

117 Lucas JM, Heinlein C, Kim T, et al. The androgen-regulated protease TMPRSS2 activates a proteolytic cascade involving components of the tumor microenvironment and promotes prostate cancer metastasis. Cancer Discovery (2014) 4(11):1310–25. doi: 10.1158/2159-8290.CD-13-1010

118 Zhou F, Gao S, Han D, et al. TMPRSS2-ERG activates NO-cGMP signaling in prostate cancer cells. Oncogene (2019) 38:4397–411. doi: 10.1038/s41388-019-0730-9

119 Ubels JL, Veenstra ED, Ditlev JA, Ingersoll KE. Retinoic Acid Inhibits Androgen-Stimulated Up-Regulation of Androgen Receptor Expression in Lacrimal Glands of Orchiectomized Rats. Invest Ophthalmol Vis Sci (2003) 44(13):1023.

120 Wambier CG, Goren A. SARS-COV-2 infection is likely to be androgen mediated. J Am Acad Dermatol (2020), S0190–9622(20)30608-3. doi: 10.1016/j.jaad.2020.04.032

121 Feily A, Taheri T, Meier-Schiesser B, Rhinehart DP, Sobhanian S, ColonDiaz M, et al. The effect of low-dose isotretinoin therapy on serum androgen levels in women with acne vulgaris. Int J Womens Dermatol (2019) 6(2):102– 4. doi: 10.1016/j.ijwd.2019.10.007

122 Dobson R. Men are more likely than women to die early. BMJ (2006) 333 (7561):220. doi: 10.1136/bmj.333.7561.220-c

123 Michelozzi P, de’Donato F, Scortichini M, et al. Mortality impacts of the coronavirus disease (COVID-19) outbreak by sex and age: rapid mortality surveillance system, Italy, 1 February to 18 April 2020. Euro Surveill (2020) 25(19):2000620. doi: 10.2807/1560-7917.ES.2020.25.19.2000620

124 Karlsson T, Vahlquist A, Kedishvili N, Törmä H. 13-cis-retinoic acid competitively inhibits 3 alpha-hydroxysteroid oxidation by retinol dehydrogenase RoDH-4: a mechanism for its anti-androgenic effects in sebaceous glands? Biochem Biophys Res Commun (2003) 303(1):273–8. doi: 10.1016/s0006-291x(03)00332-2

125 Wu C, Liu Y, Yang Y, Zhang P, Zhong W, Wang Y, et al. Analysis of therapeutic targets for SARS-CoV-2 and discovery of potential drugs by computational methods. Acta Pharm Sin B (2020) 10(5):766–88. doi: 10.1016

126 Esteban L, Eisman J, Gardiner E. Vitamin D receptor promoter and regulation of receptor expression. In: Feldman D, Pike JW, Glorieux FH, editors. Vitamin D. 2. New York: Elsevier/Academic Press; 2005. pp. 193– 217

127 Cheng TY, Goodman GE, Thornquist MD, Barnett MJ, Beresford SA, LaCroix AZ, Zheng Y, Neuhouser ML. Estimated intake of vitamin D and its interaction with vitamin A on lung cancer risk among smokers. Int J Cancer. 2014 Nov 1;135(9):2135–45. doi: 10.1002/ijc.28846. Epub 2014 Mar 24; PMID: 24622914; PMCID: PMC4293152..

128 Aranow C. Vitamin D and the immune system. J Investig Med. 2011;59(6):881C:886.

129 Khoo AL, Chai L, Koenen H, Joosten I, Netea M, van der Ven A. Translating the role of vitamin D3 in infectious diseases. Crit Rev Microbiol. 2012;38(2):122C:135

130 Anthony R. Mawson, “Role of Fat-Soluble Vitamins A and D in the Pathogenesis of Influenza: A New Perspective”, International Scholarly Research Notices, vol. 2013, Article ID 246737, 26 pages, 2013. https://doi.org/10.5402/2013/246737.

131 Demir M, Demir F, Aygun H. Vitamin D deficiency is associated with COVID-19 positivity and severity of the disease. J Med Virol. 2021 May;93(5):2992–2999. doi: 10.1002/jmv.26832. Epub 2021 Feb 9; PMID: 33512007; PMCID: PMC8013436.

132 Karahan S, Katkat F. Impact of Serum 25 (OH) Vitamin D level on mortality in patients with COVIDC:19 in Turkey. J Nutr Health Aging. 2020:1C:8

129 Entrenas Castillo M, Entrenas Costa LM, Vaquero Barrios JM, et al. Effect of calcifediol treatment and best available therapy versus best available therapy on intensive care unit admission and mortality among patients hospitalized for COVIDC:19: A pilot randomized clinical study. J Steroid Biochem Mol Biol. 2020;203:105751.

133 Entrenas Castillo M, Entrenas Costa LM, Vaquero Barrios JM, et al. Effect of calcifediol treatment and best available therapy versus best available therapy on intensive care unit admission and mortality among patients hospitalized for COVIDC:19: A pilot randomized clinical study. J Steroid Biochem Mol Biol. 2020;203:105751

134 Tepasse, P. R., Vollenberg, R., Fobker, M., Kabar, I., Schmidt, H., Meier, J. A., Nowacki, T., & Hüsing-Kabar, A. (2021). Vitamin A Plasma Levels in COVID-19 Patients: A Prospective Multicenter Study and Hypothesis. Nutrients, 13(7), 2173. https://doi.org/10.3390/nu13072173

135 Kartal, D., Yaşar, M., Kartal, L., Özcan, I., & Borlu, M. (2017). Effects of isotretinoin on the olfactory function in patients with acne. Anais brasileiros de dermatologia, 92(2), 191–195. https://doi.org/10.1590/abd1806-4841.20175483.

136 Midha IK, Kumar N, Kumar A, Madan T. Mega doses of retinol: A possible immunomodulation in Covid-19 illness in resource-limited settings. Rev Med Virol. 2021 Sep;31(5):1–14. doi: 10.1002/rmv.2204. Epub 2020 Dec 31; PMID: 33382930; PMCID: PMC7883262.

137 Bahmad HF, Abou-Kheir W. Crosstalk between COVID-19 and prostate cancer. Prostate Cancer Prostatic Dis. 2020 Dec;23(4):561–563. doi: 10.1038/s41391-020-0262-y. Epub 2020 Jul 24; PMID: 32709978; PMCID: PMC7378980.

138 Saini, K.S., Lanza, C., Romano, M. et al. Repurposing anticancer drugs for COVID-19-induced inflammation, immune dysfunction, and coagulopathy. Br J Cancer 123, 694–697 (2020). https://doi.org/10.1038/s41416-020-0948-x

139 Cadegiani, F.A. Repurposing existing drugs for COVID-19: an endocrinology perspective. BMC Endocr Disord 20, 149 (2020). https://doi.org/10.1186/s12902-020-00626-0

140 Kouhpayeh S, Shariati L, Boshtam M, Rahimmanesh I, Mirian M, Esmaeili Y, Najaflu M, Khanahmad N, Zeinalian M, Trovato M, Tay FR, Khanahmad H, Makvandi P. The Molecular Basis of COVID-19 Pathogenesis, Conventional and Nanomedicine Therapy. Int J Mol Sci. 2021 May 21;22(11):5438. doi: 10.3390/ijms22115438; PMID: 34064039; PMCID: PMC8196740.

141 Krumm, Z.A., Lloyd, G.M., Francis, C.P. et al. Precision therapeutic targets for COVID-19. Virol J 18, 66 (2021). https://doi.org/10.1186/s12985-021-01526-y

142 Bravaccini S, Fonzi E, Tebaldi M, Angeli D, Martinelli G, Nicolini F, Parrella P, Mazza M. Estrogen and Androgen Receptor Inhibitors: Unexpected Allies in the Fight Against COVID-19. Cell Transplant. 2021 Jan-Dec;30:963689721991477. doi: 10.1177/0963689721991477; PMID: 33522308; PMCID: PMC7863556.

143 Shoemark DK, Colenso CK, Toelzer C, Gupta K, Sessions RB, Davidson AD, Berger I, Schaffitzel C, Spencer J, Mulholland AJ. Molecular Simulations suggest Vitamins, Retinoids and Steroids as Ligands of the Free Fatty Acid Pocket of the SARS-CoV-2 Spike Protein*. Angew Chem Int Ed Engl. 2021 Mar 22;60(13):7098–7110. doi: 10.1002/anie.202015639. Epub 2021 Feb 22; PMID: 33469977; PMCID: PMC8013358.

144 Hakim, A. S., Syame, S. M., Shehata, M. M., & Sayed, A. T. A. (2020). The Battle with COVID-19: Insight on External Intervention and Future Vaccination. South Asian Journal of Research in Microbiology, 7(2), 46–61. https://doi.org/10.9734/sajrm/2020/v7i230169

145 Barh D, Tiwari S, Weener ME, Azevedo V, Góes-Neto A, Gromiha MM, Ghosh P. Multi-omics-based identification of SARS-CoV-2 infection biology and candidate drugs against COVID-19. Comput Biol Med. 2020 Nov;126:104051. doi: 10.1016/j.compbiomed.2020.104051. Epub 2020 Oct 10; PMID: 33131530; PMCID: PMC7547373.

